# Sterilization of disposable face masks with respect to COVID-19 shortages; a nationwide field study including 19 sterilisation departments

**DOI:** 10.1101/2021.01.15.21249843

**Authors:** B. van Straten, P.D. Robertson, H. Oussoren, S. Pereira Espindola, E. Ghanbari, J. Dankelman, S. Picken, T. Horeman

**Author notes:** **Author for correspondence:** Name: Bart van Straten, Department: Department of BioMechanical Engineering, Institution: Delft University of Technology, Address: Mekelweg 2, Building 34, 2628 CD Delft, The Netherlands.

## Abstract

**Objective:** Face masks also referred to as half masks are essential to protect healthcare professionals, working in close contact with patients having Covid-19 related symptoms. During the threating deficit, healthcare institutions sought an approach to re-use face masks or to acquire imported masks. The objective of this study is to assess the quality of sterilised and imported FFP2/KN95 face mask materials.

**Design:** prospective, bench-to-bedside

**Setting:** General healthcare including 19 hospitals in the Netherlands

**Interventions:** Face masks were reprocessed using a medical autoclave at 121°C.

**Methods:** A 48 minutes steam sterilization process of single-use face masks with 15 min holding time at a 121 °C was developed, validated and implemented in 19 different hospitals. Steam and H_2_O_2_ plasma sterilized as well as new, imported masks are tested in a custom-made, non-standard EN-149, test set-up that measures Particle Filtration Efficiency (PFE) and pressure drops.

**Results:** PFE validation data of 84 masks indicated differences of 2.3±2 % (mean±SD) between the custom build test set-up and a continues flow according to the EN-149. Test data showed the mean PFE values of 444 sterilised FFP2 face masks from 19 CSSD were 90±11% (mean±SD) and of 474 imported KN95/FFP2 face masks 83±16% (mean±SD). Differences in PFE between sterilisation departments were found.

**Conclusion:** Face masks can be reprocessed with 121 0 C steam or H_2_O_2_ plasma sterilization with minimum reduction of PFE. PFE comparison between sterilised mask and new, imported mask filter material indicates that most reprocessed masks of high quality brands outperform new imported face masks of unknown brands. Although the PFE of tested face mask material from different sterilisation departments remained efficient, different types of sterilisation equipment can result in different PFE outcomes.

**Strengths and limitations of this study:**

– Reprocessing face masks at 121 °C steam Sterilization, a simple method to be used by hospitals in times of shortages
– Laboratory findings to evaluate the safety and quality of face mask material
– The study is limited and restricted to selected FFP-2 face masks
– This study is a first of its kind in quality and safety check of the vast growing face masks, entering our markets
– The study focusses on testing environmental dry particles in a rapid test setup

## INTRODUCTION

After the outbreak of Covid-19, this respiratory disease has been spreading in a highly rapid pace [1,2]. Adequate face masks are essential to protect healthcare professionals. In many hospitals shortages of personal protection equipment arose due to increased demands [3]. In the search for alternative sources, hospitals started to consider reuse by sterilizing single-use face masks [4].

Face masks, also referred to as half masks, are used during aerosol generating procedures to protect against airborne particles. Three classes of Filtering Facepiece Particle (FFP) are described in the European Norm (EN) 149:2001+ A1:2009 [5]. The most used one in relation to Covid-19 are the Class 2 FFP2 masks which are considered to be equivalent to the American N95 [6] conforming the standards of the National Institute for Occupational Safety and Health (NIOSH) 42 CFR 84 [7] and the Chinese KN95 complying to the Guobiao (GB) 2626-2006 standard. [8] The filter efficiency of the smaller particles is a crucial element. The European Norm (EN) requires a minimum filter efficiency of 94% whereas NIOSH [7] and GB [8] require 95%.

### Testing filter material of face mask

The EN 149:2001+A1:2009 [5] and more specific NEN-EN 13274-7:2019, part 7 describes a test setup that consists of a flow tube, a flow generator, a NaCl particle generator and two particle measurement devices to determine the Particle Filtration Efficiency (PFE) of face masks with different flows up to 120 l/min and NaCl particles of 0.1 to 10 µm. Unfortunately, this setup is costly to build. Therefore, in the first 2 months of Covid-19, only 2 systems were operational in the Netherlands and used for testing of new, imported face masks. The costs of tests of one face mask was around 1,500 Euro with a waiting list of up to 4 weeks. A new quick testing method was needed.

### Potential reprocessing methods

Multiple studies show the effect of different sterilization methods including Gamma sterilization, Plasma sterilization, steam and dry heat sterilization, microwave, washing machine and UV–C light as methods to decontaminate face masks for the purpose of reuse [9-13]. These studies suggest that Gamma, and steam sterilization conducted at 134 °C damage the micro structure of the filter material [9].

Washing machine and microwave have a low capacity and the microwave does not create a uniform heat distribution and requires a steam bag [10-12]. Some studies suggest that the high concentration of liquid H_2_O_2_ in plasma sterilization (approx. 60%) and its strongly charged ionized vaper may neutralize filter media’s electrostatic charge [11,12]. Moreover, the sterilization’s efficacy would likely be affected by presence of moisture (e.g. exhaled breath) in worn masks, as water is a polar molecule. Finally, the capacity per run remains low due to the vacuum driven process [13] and the evaporation of moisture may restrict the sterilizer’s ability to pull deep vacuum. UV treatment of face masks seems to have potential but requires preparation time as face masks need to be unfolded such that the UV light reaches all the mask material [10-13]. Steam sterilization at 121 °C could be an option since studies have shown effectiveness at 121 °C to inactivate the coronavirus [14,15].

Since hospitals needed to know if 121 °C sterilisation method was safe and effective, pilot studies were conducted that included ATCC 12228 bacteria testing. After demonstrating that sterilisation up to 5 times is possible for high quality face masks, we made the protocols and results available to hospitals via the repository of the Delft University of Technology [9,16]. However, the accuracy of this new method was not explored. Moreover, a study of many different brands processed at different CSSDs with comparison between new, imported masks and sterilised masks do not exist.

The aim of this study is to assess the quality of FFP2 face masks after H_2_O_2_ plasma and steam sterilization.

The following research questions were defined:

1. Can FFP2 masks be reprocessed using 121 °C steam or H_2_O_2_ plasma sterilization?
2. Are reprocessed face masks an alternative for new ones?
3. What effect does sterilization have on the materials?

## METHODS

A sterilization facility of a Dutch CSSD (ISO 7 validated, Van Straten Medical, De Meern, the Netherlands, operated by CSA services) was rebuilt for the purpose of reprocessing used (potential Covid-19 contaminated) FFP2 face masks. New testing methods were built to test the filter material quality after sterilization [4,9,16]. The testing facility was open for any hospital, reseller, manufacturer to check the quality of sterilized or new face masks.

### Reprocessing by 121 °C steam at CSA Services Sterilization

To implement the 121 °C sterilization process, a separate logistical routing was made for collecting and processing used face masks. Upon receipt the masks were taken out of the double wrapping and inspected individually for visual damage. In case of deformities or dirt, lipstick, hairs, black streaks, stains and other deviations, the masks were discarded. The visually approved masks were marked with a dot and packaged in autoclavable impermeable sterilization laminate bags (type CLFP150×300WI-S20, Halyard, UK) (Figure 1). After a mask was marked with the maximum of 5 dots it was discarded. A maximum of five face masks were packaged per bag in order to have them sterilized properly. The autoclaves (GSS6713H-E, Getinge, Sweden) were activated with a 121 °C program and re-validated. The autoclave cycle was set on 48 minutes with a 15 min holding time (high vacuum 121 °C; ≥15 min HT, total CT 48 min). Face masks with a higher class (FFP3/N95) were treated as FFP2/KN95. The PFE for the average particle size of a FFP2/KN95 mask material should be 94 % or higher for a pass, and under 94 % for a fail [5]. The performance of the masks material was determined by measuring the PFE and breathing potential. Figure 2 shows the particle counter with a custom-made particle chamber connected (Lighthouse Solair 3100, San Francisco, www.golighthouse.com). The machine draws air from the surrounding, through the mask into the chamber and tube, to the particle counter to provide a number of particles per size. The diameter of the chamber is chose such that it guarantees sufficient airflow through the filter material to match the specifications of the particle counter device [9,16,17]. The PFE was determined by measuring the difference in number of particles before and after filtration by the mask. First, the particle concentration in a standard volume of room air was determined by measuring the number of particles (sizes 0.3, 0.5, 1.0, and 5.0 μm) in a volume of surrounding air. Second, the mask was installed on the chamber to measure the number of particles after filtration.

**Figure 1.**
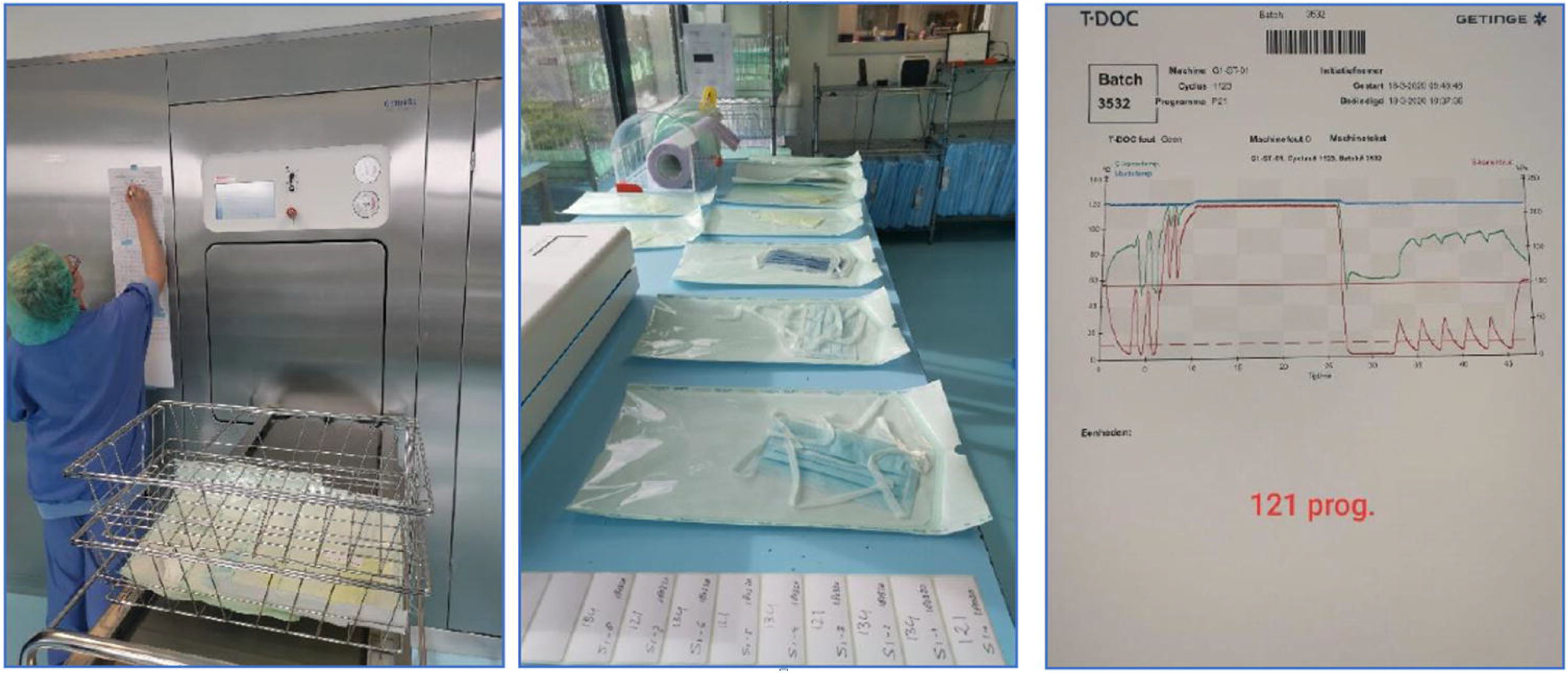
Autoclave procedure with Halyard laminate bags. Left, laminated bags entering the autoclave. Middle, masks are wrapped in laminate. Right, the 121 °C steam sterilisation program as used for face mask sterilisation.

**Figure 2.**
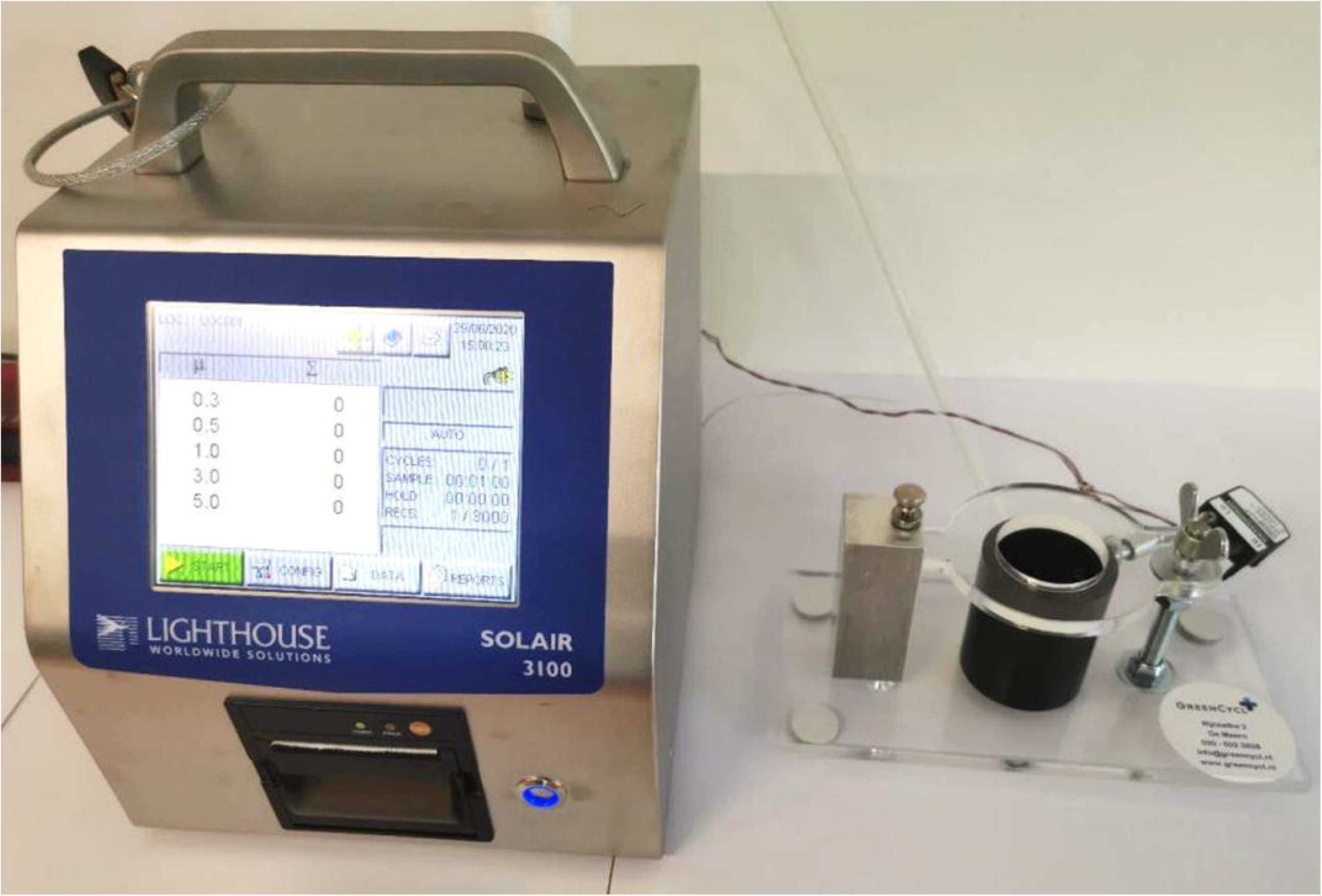
Lighthouse Solair 3100 particle counter connected to a particle chamber.

When very dense filter materials are used, with a very high PFE for the smallest particles, it causes a breathing resistance for the user [16,17]. This breathing potential was determined by measuring the pressure drop using an analog differential pressure sensor, type SDP2000-L connected to the particle chamber. The pressure sensor is temperature compensated, calibrated and has a resolution of 11 Pa with a repeatability of 0.3 % and accuracy of 1 % [17]. The breathability requirements for respiratory protective devices are provided in a European standard [18]. The maximum permitted resistance (mbar) differs for FF1, FFP2, and FFP3 masks, ranging from 0.6-1.0 for inhalation at 30 l/min, 2.1-3.0 for 95 l/min and is 3.0 for exhalation at 160 l/m. The norm for a FFP2-mask at 30 l/min is 0.7 mbar.

### Test Setup validation to European Norm

The accuracy of the developed particle test setup was evaluated by comparing results from known face masks, tested on (our) particle setup, with the results of the same brand and type masks, tested on a continuous flow system. The continuous flow tests system used NaCl particles and was built at the Delft university of technology according to the NEN-EN 13274-7:2019 [19]. The EN-149 standard of Table I includes experiments to determine the inward leakage. Therefore, a Fit-test and strap-test may be conducted, conforming a proper fit on the face without leakages around the mask [17,18]. In this study, inspection of the materials and leakage test were conducted on all reprocessed masks. Although we focussed on the material properties in this study, only masks that showed no change in fit or material properties were included the types that did deteriorate were registered and disposed after arrival. Although we followed the EN-149 norm as much as possible we did reference our outcomes with the NaCl test since we used a custom made test setup as a non-standard EN-149 methodology.

**Table I.** EN 149 tests.

| FFP 2 EN-149 Criteria | % |
| --- | --- |
| Filter airborne particles, at least | 94 |
| <u>Inward leakage of up to</u> | 8 |
| EN 13274-7: European Standard for testing particle filter penetration, 2 Aerosol tests: | NaCl test,<br>Paraffin oil |

### 121 °C steam sterilisation consistency between CSSDs

The consistency of sterilisation results caused by different processes and equipment was compared between 19 CSSDs. Samples of masks representing most commonly used brands and types were selected and measured with the PFE setup. Only CSSDs were included that provided minimal four masks that were sterilised only once. Face masks were not cleaned after visual inspection and prior to sterilization. A Student T-test (two tailed, unequal variance, SPSS 17.0) was used for comparison and a probability of p<0.05 was considered to be statistically significant.

### Face mask material differences

Differences in mask material can be analysed by chemically and thermally comparing the fabric of the two most common types, showing different PFE after being sterilised with 121 °C or H_2_O_2_ plasma sterilisation. Therefore a Differential Scanning Calorimetry (DSC), X-Ray Diffraction (XRD) and Transform InfraRed spectroscopy (FTIR) were conducted (Supplemental file 1).

### Testing new masks

Samples were selected for PFE measurement from batches of imported masks. The PFE results of those new face masks were compared to the PFE results of the sterilized face masks from the 19 CSSDs. New imported face masks that scored above 98 % PFE in the particle range were further investigated by measuring the pressure drop.

## RESULTS

### Reprocessing by 121 °C steam sterilization at CSA services

74,834 masks from hospitals were processed by CSA Services. Of these masks 56,668 were disposed after incoming inspection due to visual damage, deformities or dirt. The remaining 18,166 face masks were steam sterilised at 121 °C. Table II shows the top 5 brands that were sterilised and released back to hospitals.

**Table II.** Top 5 reprocessed face masks.

| **Brand (Type)** | **Percentage** |
| --- | --- |
| 3M (1862+) | 42% |
| 3M (1872+) | 21% |
| My-T-Gear | 8% |
| IMG Europe (R620) | 5% |
| Kimberly Clarc Corp | 5% |
| Rest | 19% |

### Test Setup validation to European Norm

Preliminary tests conducted with 84 different masks tested on PFE dry particle test setup and a NaCl test setup built according to the NEN-149 indicated an outcome deviation of 2.3 (SD 2) % on average with a max of 7 % (supplemental file 2). A measurement test conducted with another 10 different masks indicated an average 19 s (SD 21) is needed to install and inspect the mask on the particle counter and an additional 15 s (SD 13) to take the mask from the system after 1 minute of measurement. None of the masks showed visual signs of deformation or damage after being measured.

### 121 °C steam sterilisation consistency between CSSDs

The reprocessing method was adopted by the CSSDs of 19 hospitals (Amsterdam University Medical Center (VUmc and AMC locations), Holendrecht Medical Center, Franciscus Hospital, CombiSter RDGG & Haga, Spaarne Hospital, Erasmuc MC, University Medical Center Groningen, Leiden University Medical Center, Flevo Hospital, Isala Hospital, Diakonessenhuis Utrecht, VieCuri, Rode Kruis Hospital, Noordwest Hospital Group, Amphia Hospital, and Tweesteden Hospital). The PFE results of 444 reprocessed FFP2/KN95 face masks from CSSDs of 19 different hospitals in the Netherlands are provided in Figure 3. From these 444 masks, 371 masks were reprocessed by steam sterilization and 73 processed by means of H_2_O_2_ plasma (supplemental file 3). From the 444 tested face masks, 58 3M 1862+ face mask were provided by seven CSSDs from four university hospitals, one general hospital and one general practitioner, only sterilised once with 121 °C steam sterilisation (supplemental file 4). The influence of different installations, protocols or staff on PFE are shown in Figure 4. The “N” value indicates how many 3M 1862+ face masks were included in the study that were only sterilised once. The statistical tests reveal differences in outcome mainly for the CSSD of University Hospital 2.

**Figure 3.**
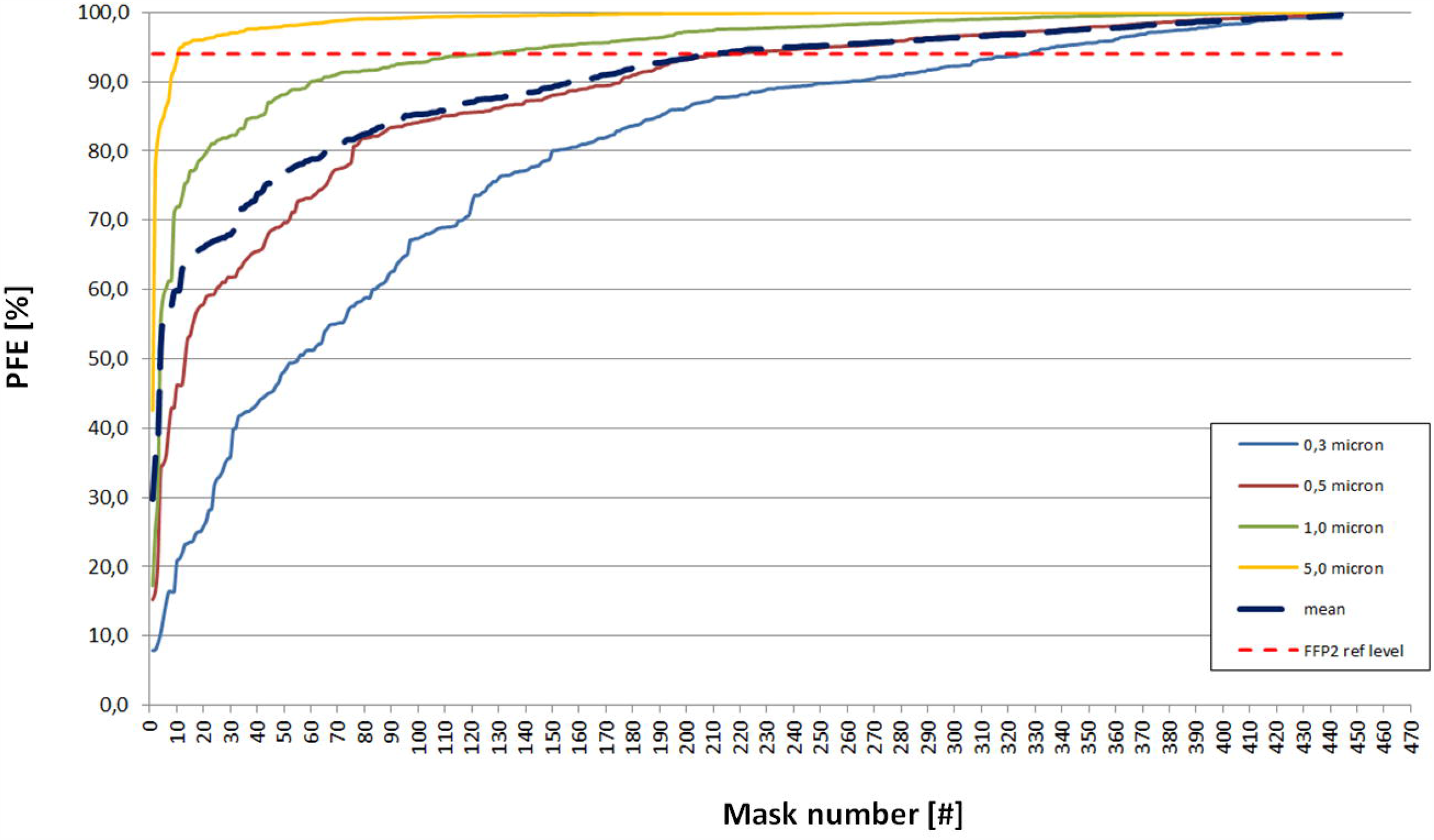
PFE values after sterilization with 121 °C steam or H_2_O_2_ plasma sterilisation in chronological order from worst to best. The red dotted line indicates the FFP2 level at 94% PFE. Each mask number represents a sample of a sterilised batch from one type only.

**Figure 4.**
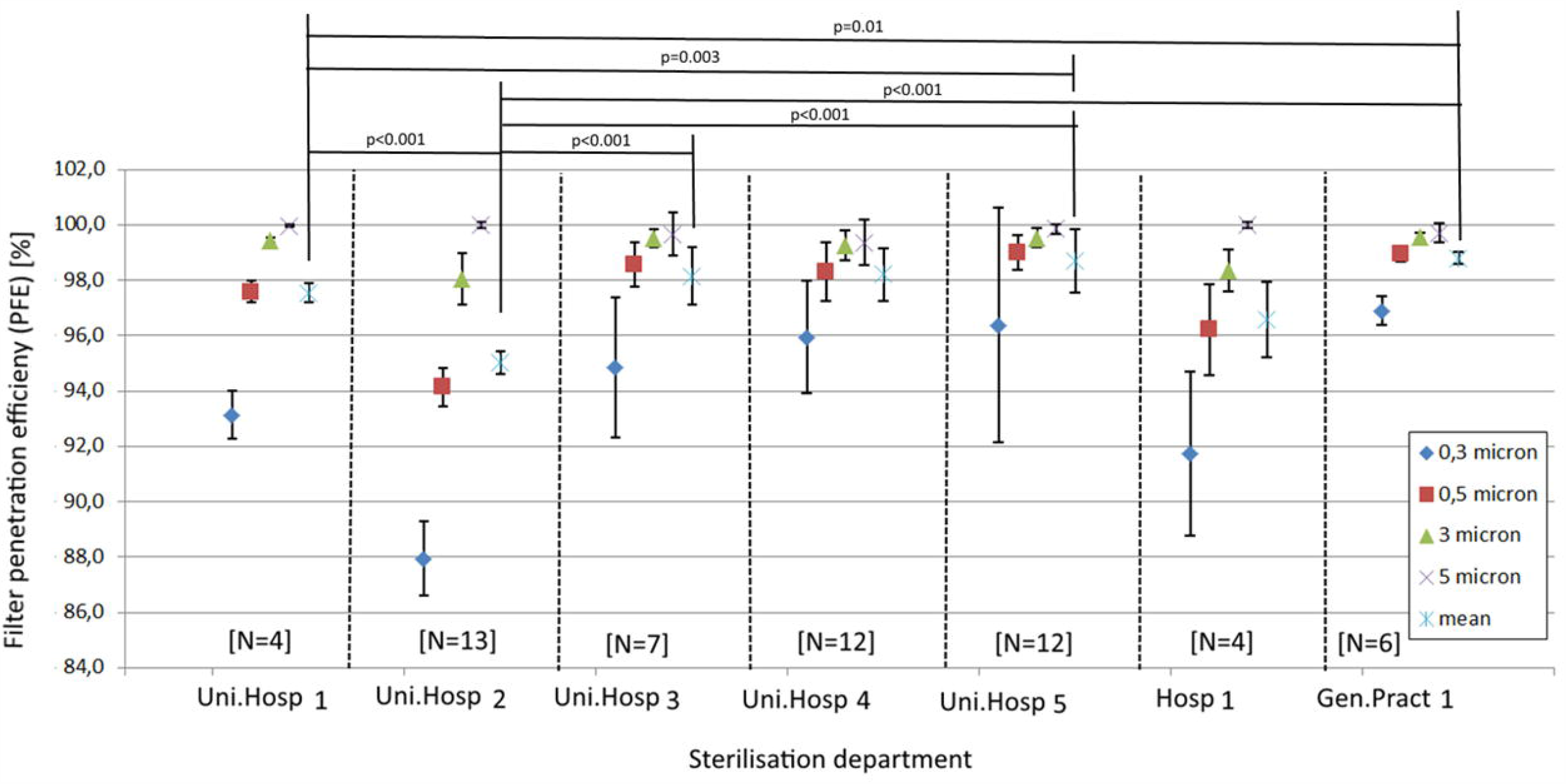
PFE values with Standard Deviation of different 3M 1862+ coming from 7 different CSSDs. Statistical differences are indicated with P values above the figure.

### Face mask material differences

The 444 masks consisted of 101 different types of masks. From the 101 different types, the 3M 1862 and Kolmi Op-Air were mostly tested. The PFE results of 89 3M 1862 and 26 Kolmi Op-Air are provided in Table III for 0.3, 0.5, 1 and 5 µm particles. The results indicate that the 3M 1862 shows low PFE values after 2x H_2_O_2_ plasma processing and Kolmi Op-Air shows low and inconsistant PFE values after 1x 121 °C processing (supplemental file 5).

**Table III.**
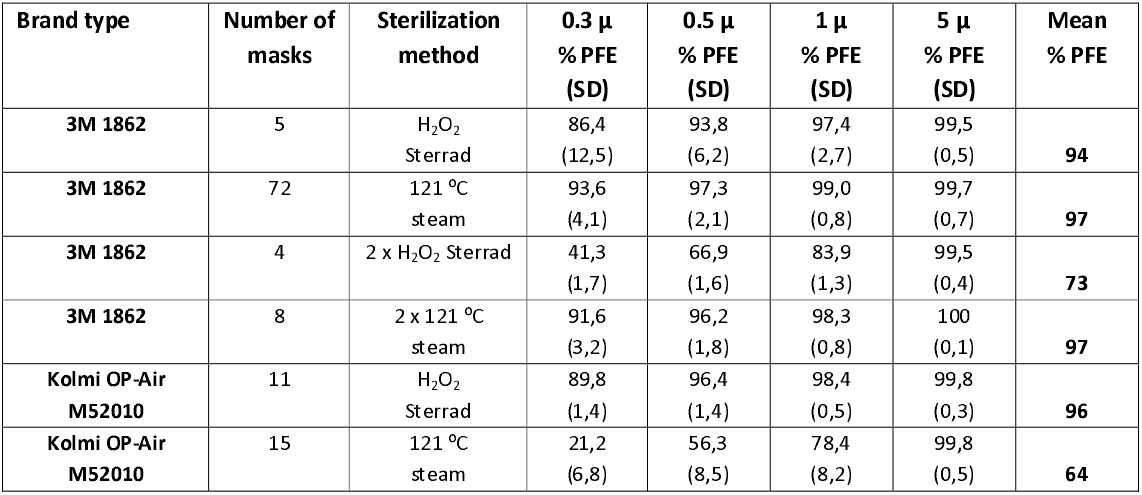
**Particle Filter Efficiency of two commonly used mask after either 121 °C steam or H_2_O_2_ Plasma sterilisation**

### Thermal properties of 3M Aura 1862+ and Kolmi Op-Air M52010 face masks using DSC

The three tests, Differential Scanning Calorimetry (DSC), X-Ray Diffraction (XRD) and Transform InfraRed spectroscopy (FTIR) confirmed a match of all 5 layers of both masks with the profile of the material PP (Supplemental file 6).

### Test new imported masks

The PFE results of 471 different types of new FFP2/KN95 imported face masks from collaborating hospitals and resellers are shown in Figure 5. From these 471 face masks, 27 face masks scored above 98 % PFE for the 0.3 micron particle size category and tested for breathability by measuring the pressure drop (supplemental file 7). Figure 6 shows the breathing potential of 27 face masks. The material of 27 face masks with high PFE values showed pressure drops between 251 and 3976 Pa on the measurement setup. When calculated for the total mask area A and B, five out of 27 masks showed a total pressure drop higher than the EU standard of 0.7 mbar [18]. Finally, 4 masks showed readings around 3.7 mbar being very close to the maximum measurable pressure drop of 4500 Pa.

**Figure 5.**
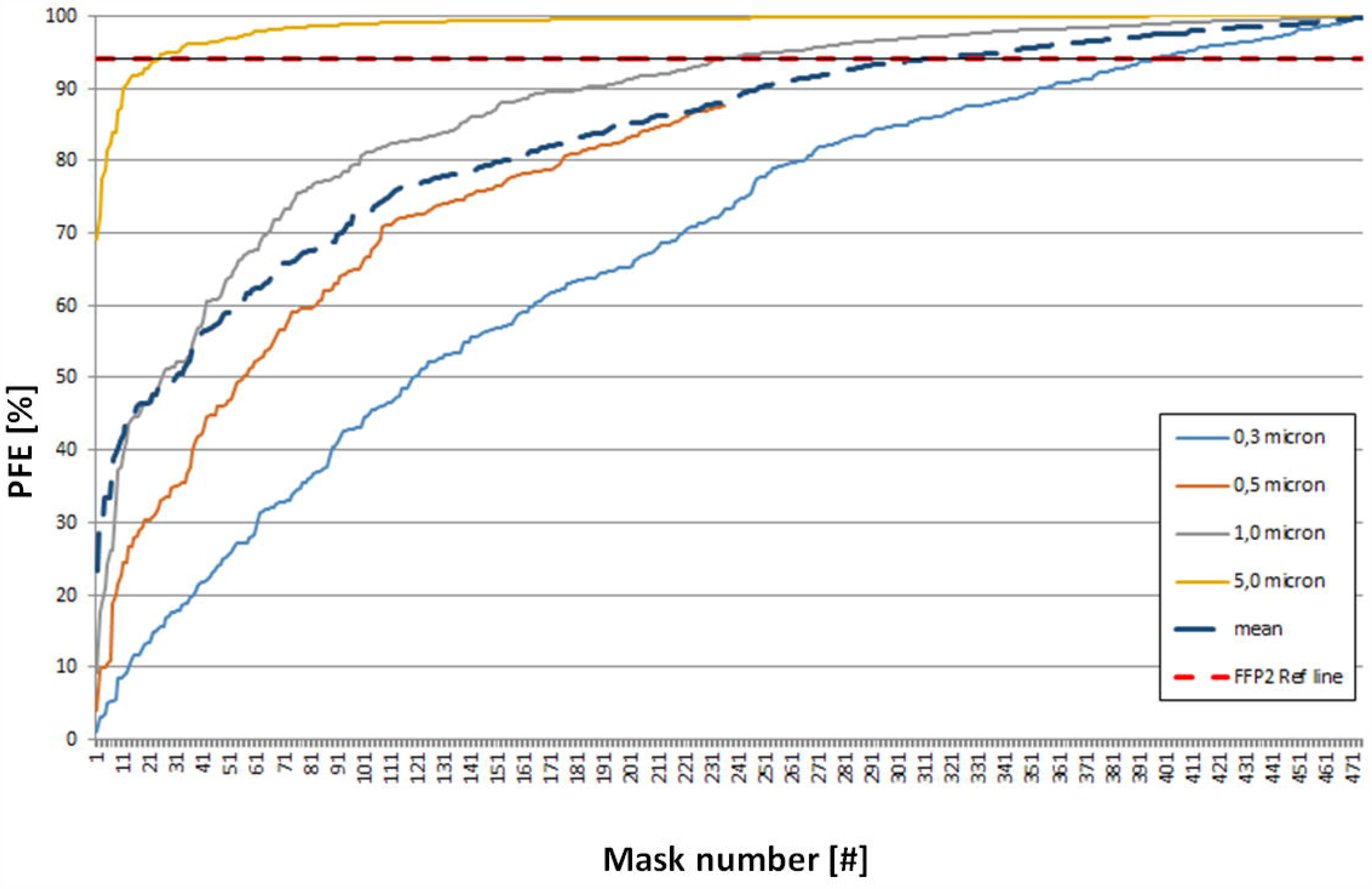
PFE values of new imported FFP2/KN95 face masks in order from worst to best. The red dotted line indicates the FFP2 level at 94% PFE. Each mask number represents a sample of a new batch from one type only.

**Figure 6.**
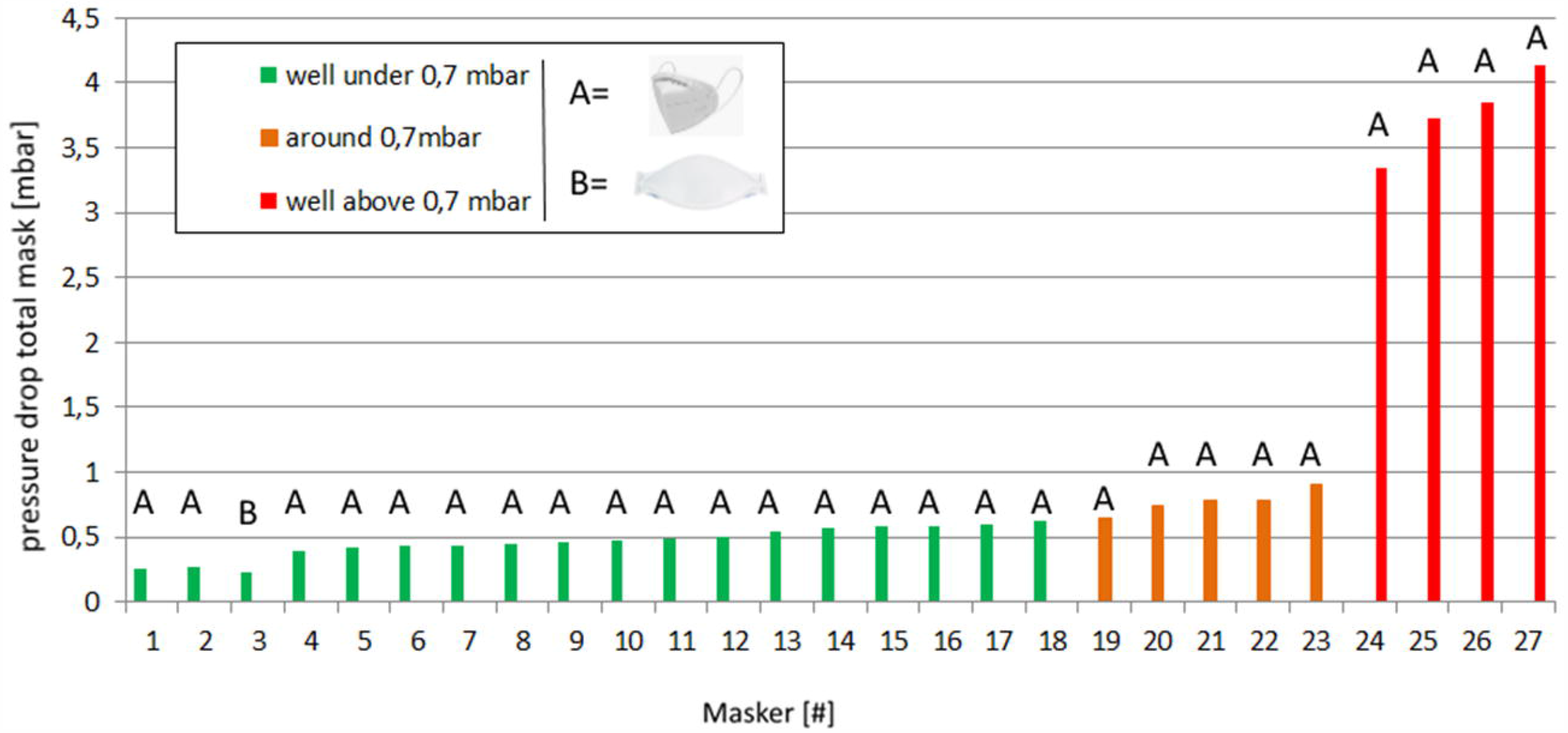
Pressure drop of 27 new face mass with PFE>0,7 mbar. Four masks performed really low (red), 5 performed around the EU norm of 0,7 mbar and 18 performed well according to the EU norm of 0,7 mbar.

## DISCUSSION

Regarding the research questions, it can be confirmed that FFP2 masks can be safely reprocessed with 121 °C steam sterilization if testing facilities are available. The data from Figures 3 and 5 indicate that reprocessed face masks can be an alternative for new face masks as sterilisation of a mask with well-known brand gives better PFE results compared to new imported masks. Although base-materials are similar, the role of manufacturing, preparation and use of coatings have a large effect on the PFE of mainly the smaller particles.

The cross validation with the NaCl continues flow setup build according to EN 13274-7, showed that the most important requirements for determining the filter material properties are met. After nineteen hospitals adapted the steam sterilization process, a nation-wide data field experiment was initiated that informed multiple international NGO’s, universities and industry members about the pros and cons of sterilisation of face masks [17-21] setting a Dutch standard for sterilisation of face masks. After the first results were shared on request [22], general practitioners, dental practices and pharmacies claimed to successfully adopted the 121 °C process in their smaller sized autoclaves with sufficient results [16].

Sterilization with the purpose of re-using medical devices is often driven by cost-savings [23]. However, some studies also report the reuse of medical devices to realize environmental benefits [24]. In this study steam sterilization is used for reasons of preventing shortages. In 1986 a survey was conducted including Canadian hospitals re-using disposable medical devices [25]. Forty-one percent of the hospitals confirmed they reused disposable medical devices with respiratory therapy equipment as most reused medical device.

Testing by particle counting seems to be essential for both new and sterilized single-use face masks since it indicates the quality of the mask in terms of filtration capacity. This is shown as our data reveals large differences in PFE despite the similar appears of the mask material. Our results in Supplement file 6 and 8 indicate the presence of coatings which improve the electrostatic behaviour of the mask. As the presence of these kind of coatings is very difficult to demonstrate, it is advised to test the PFE with a particle counter at all times. To rule out that reprocessed as well as new face masks are not meeting the stated FFP standard, a particle test as a ‘quick and dirty test’ could be applied on every batch. Therefore the test method as described in this study will lead to a quick indication of the quality.

Face masks sterilized with the intention to reuse could furthermore, undergo a “Fit test”. This test may be regarded as a fit validation conforming a proper fit on the face without leakages around the mask. In order to assure a decreased risk of spreading other diseases, also the bio efficacy of a face mask should be considered. Tests regarding this aspect were conducted previously and appeared negative for bacteria’s on steam sterilised face masks which were tested at the dept. of Microbiology at the Franciscus Hospital in the Netherlands [9].

Testing face masks on particles is important to quality assure the sterilization process as our data shows that despite the implementation of similar 121 °C sterilisation protocols, mean PFE outcomes can differ up to 6%. As the type of mask and sterilisation methods are similar, the only unknown variable is the wearing/processing-influence on the mask during use, transport and inspection. University Hospital 2 in Figure 4 seems to show a much lower PFE outcomes. It could be that stretching and bending of the mask can influence the integrity. However, is also expected that the confidence interval would have been larger as the intensity of the stretching and bending is human dependent. As the confidence interval of the mean PFE outcomes of University Hospital 2 seems similar or even smaller compared to other hospitals, it is advisable to do a validation tests that include all CSSDs.

With the CSSD at De Meern, a 10 % tolerance was accepted for sterilised sterilized face masks after testing, therefore a level of 84% filtration capacity on a 0.3 μm particle level was the minimum limit. Although not based on any evidence in literature, this percentage was considered to be sufficient with respect to the shortages of face masks taking into consideration that the Coronavirus (SARS-CoV-2) is mainly spread through 0.3 μm or larger droplets. However, a consensus needs to be made in order to actually define the minimal allowable PFE values in times of crisis.

The DSC, XRD and FTIR tests results in Supplemental file 6 conducted on each of the five layers of the 3M Aura 1862+ and Kolmi op-Air M52010 mask reveal that all layers are made from the same Polypropylene material. The differences in behavior when sterilized cannot be explained from chemical composition perspective. A detailed interpretation of the results can be found in Supplemental file 8.

The data of 410 sterilised and 471 new imported KN/N95 or FFP2 Face masks reveal that despite the differences in PFE reduction between different sterilisation process, roughly 75% of the face masks of known brands still reaches the FFP2 standard after sterilisation when compared to only 50% of new imported, less common brands. Our results suggests that the technology needed to manufacture a good mask is not easy and manufacturing and quality assurance should be monitored and controlled by the government. During the study period it was observed multiple times that within a single batch of imported face masks, the quality and layout of the masks was different despite the often clearly visual PFE standard printed on the box. In addition, some masks (Figure 6) showed almost complete lack of air penetration.

Although most users indicated to use sterilized face masks over unknown, new masks with unknown filter efficiency, expecting healthcare workers to wear masks of others can have an psychological impact. To overcome this issue masks can be marked with the initials of the user in order to return it to the same person.

### Study limitations

It is of utmost importance that the reprocessing of single use PPE, such as described in this study, are in equivalence with existing standards. Each deviation or omission of such standards need a clear demonstrated equivalence with the applying standards. In our setup, solely environmental dry particles were used in the developed rapid test setup. Although we validated the dry particle setup with an aerosol testing setup (NaCl test, Paraffin oil setup) build according to the EN 149, it was only possible to compare the PFE for a limited range of particle sizes. Therefore, in depth knowledge about the PFE related to particle size was not generated. In order to identify potential other differences between the dry particle and continues flow setups a “gap” analysis should be conducted. Other than testing the basic material of the filter layers, we were not able to indicate the presence of surface active coatings. Therefore it was not possible to investigate the role of surface active coatings on the melting or oxidation of the fibres.

## CONCLUSION

Sterilization of disposable face masks by means of standardized steam sterilization on 121 °C could be an alternative against face mask shortages due to Covid-19 if the Fit does not change. The 121 °C sterilization process can be safely implemented in different sterilisation departments as the 6 different installations show acceptable PFE results. The new PFE testing method proved to be accurate in order to determine degeneration of the mask material after sterilization and to determine the material quality of imported face masks. As FTIR, XRD and DSC indicate that all layers from both masks are made from PP, differences in

PFE outcome after being sterilised with 121 °C steam or H_2_O_2_ plasma sterilisation can be explained by the level of crystallinity and the orientation and dimensions of the fibres and potential proprietary treatment in the layers of the face mask. PFE comparison between sterilised masks and imported face masks with varying filter qualities, indicates that health care professionals in some cases can better reuse a known reprocessed brand rather than an imported face mask from a reseller with an unknown brand.

## Data Availability

Technical appendix
All data is included in supplemental files.

## Acknowledgment

The authors like to thank the CSSD personnel and sterilisation specialists from all participating hospitals for sending us samples of their sterilised batches for testing. Special thanks to the personnel and management from CSA Services for transforming their CSSD into a high volume face mask processing line. The students and colleagues of TU-Delft that helped testing during “ProjectMask” are thanked for their effort. Finally, resellers are thanked for testing their masks at our test facility in De Meern and Delft University of technology.

## Author statement

### Contributors

B. van Straten, J. Dankelman, T. Horeman contributed to the study design and acquisition of data. P.D. Robertson, H. Oussoren, S. Pereira Espindola, E. Ghanbarim S. Picken analyzed and interpreted the data. B. van Straten and T. Horeman drafted the initial manuscript and all authors critically revised the manuscript and gave final approval.

## Technical appendix

All data is included in supplemental files.

Patient consent for publication

Not required.

## Ethics approval

Not required.

## References

1 Oberfeld B, Achanta A, Carpenter K, Chen P, Gilette N, Langat P & Pillai, S. SnapShot: COVID-19. Cell 2020.

2 ROSALES-MENDOZA, Sergio, et al. What does plant-based vaccine technology offer to the fight against COVID-19?. Vaccines, 2020, 8.2: 183.

3 ADDI, Rachid Ait; BENKSIM, Abdelhafid; CHERKAOUI, Mohamed. Easybreath Decathlon Mask: An Efficient Personal Protective Equipment (PPE) against COVID-19 in Africa. Journal of Clinical & Experimental Investigations/Klinik ve Deneysel Arastirmalar Dergisi, 2020, 11.3.

4 Van Den Dobbelsteen J. J., Van Straten B. J., Horeman T. A Comparison of Particle Filter Efficiency Measurements for Protective Masks using Particle Counters with Different Flow Rates. 2020.

5 European Committee for Standardization. CEN/TC 79 - Respiratory protective devices 2009.

6 Lee SA, Hwang DC, Li HY, Tsai CF, Chen CW, Chen JK. Particle Size-Selective Assessment of Protection of European Standard FFP Respirators and Surgical Masks against Particles-Tested with Human Subjects. J Healthc Eng. 2016; 8572493. doi:10.1155/2016/8572493.

7 CDC. The National Institute for Occupational Safety and Health (NIOSH) 2014 Retrieved from https://www.cdc.gov/niosh/docs/96-101/default.html

8 Code of China. Chinese Standard GB 2626-2006. Respiratory protective equipment – China Safety 2006. Retrieved from: https://www.codeofchina.com/standard/GB2626-2006.html

9 de Man, P., van Straten, B., van den Dobbelsteen, J., van der Eijk, A., Horeman, T., & Koeleman, H. (2020). Sterilization of disposable face masks by means of standardized dry and steam sterilization processes; an alternative in the fight against mask shortages due to COVID-19. Journal of Hospital Infection, 2020, 105(2), 356–357.

10 Schöpe H. J., & Klopotek M. Strategies for the re-use of FFP3 respiratory masks during the COVID-19 pandemic 2020. arXiv preprint arXiv:2004.00769.

11 Dennis J. Viscusi, Michael S. Bergman, Benjamin C. Eimer and Ronald E. Shaffer. Evaluation of Five Decontamination Methods for Filtering Facepiece Respirators, Annals of Occupational Hygiene 2009; 53, 815–827

12 Michael S. Bergman et al. Evaluation of Multiple (3-Cycle) Decontamination Processing for Filtering Facepiece Respirators, Journal of Engineered Fibers and Fabrics 2010; 5,33–41

13 Kumar A, Kasloff S. B, Leung A, Cutts T, Strong J.E., Hills K, Krishnan, J. N95 mask decontamination using standard hospital sterilization technologies. medRxiv 2020.

14 Henwood T. Coronavirus disinfection in histopathology. Journal of Histotechnology 2020; 1–3. 10.1080/01478885.2020.1734718.

15 Zhang Qinxin & Zhao, Qingshun. Inactivating porcine coronavirus before nuclei acid isolation with the temperature higher than 56 °C damages its genome integrity seriously 2020.

16 Harskamp R.E, Van Straten B, Bouman J, Van Maltha-Van Santvoort B, Van den Dobbelsteen J. J., Van der Sijp J.R., Horeman T. Reprocessing filtering facepiece respirators in primary care using medical autoclave: prospective, bench-to-bedside, single-centre study. BMJ open 2020; 10(8), e039454.

17 Teesing G.R., van Straten B, de Man P, & Horeman-Franse T. Is there an adequate alternative to commercially manufactured face masks? A comparison of various materials and forms. Journal of Hospital Infection 2020; 106(2), 246–253.

18 European Committee for Standardization. NEN-EN 149+A1 (en): Respiratory protective devices - Filtering half masks to protect against particles - Requirements, testing, marking 2009.

19 National Institute for Public Health and the Environment, RIVM. Reprocessing FFP-type masks 2020; Data retrieved from https://www.rivm.nl/coronavirus-covid-19/professionals/binnen-ziekenhuis/mondmaskers

20 3M. Decontamination of 3M Filtering Facepiece Respirators: Global Considerations, technical Bulletin, revision3 2020; https://multimedia.3m.com/mws/media/1851918O/decontamination-of-3m-filtering-facepiece-respirators-global-considerations.pdf

21 Markus A, Contingency Reprocessing of Single-Use Personal Protective Equipment (PPE), Issue 2 White paper. Retrieved from: https://www.belimed.com/en-us/success-stories/case-studies.

22 Van Straten B, Van den Dobbelsteen J, Horeman T. Steam sterilization of used disposable face masks with respect to COVID-19 shortages 2020; http://resolver.tudelft.nl/uuid:078a3733-84d6-4d4a-81e6-74210c7fed78

23 Panta, Gopal & Richardson, Ann & Shaw, Ian. Effectiveness of autoclaving in sterilizing reusable medical devices in healthcare facilities. The Journal of Infection in Developing Countries 2019; 13. 858–864. 10.3855/jidc.11433.

24 van Straten, B., Dankelman, J., van der Eijk, A., & Horeman, T. (2020). A Circular Healthcare Economy; a feasibility study to reduce surgical stainless steel waste. Sustainable Production and Consumption, 27, 169–175.

25 Campbell B.A. & Wells, G.A. & Palmer, W & Martin D. Reuse of disposable medical devices in Canadian hospitals. American journal of infection control 1987; 15. 196–200. 10.1016/0196-6553(87)90095-2.

