## Supplemental file 8 for "Sterilization of disposable face masks with respect to COVID-19 shortages; a nationwide field study including 19 sterilisation departments"

Supplemental file 8 Discussion material test results

The XRD tests reveal that for 3M 1862+ face mask, the main peak positions are in good agreement with known PP data [1,2] as PP crystallizes in a monoclinic unit cell with a = 6.630, b = 20.780, c = 6.500 Å and β = 99.0 degrees, it shows 2teta (miller indices, hkl) peaks at 16.5 (110), 19.8 (040), 21.7 (130), 24.9 (111), 25.5 (131), 30.0 (060) and 33.3 (220) degrees. Layer 1,2,3 can be defined as PP, which is the commonly used non-woven fabric material for face masks. The peak positions of Layer 4 are also in line with the PP data. However, the peaks at 24.9 (111) and 25.5 (131) are not clearly observed. 3M-4 and 3M-5 layers are considered as PP and the missed peaks maybe caused by orientation during fiber spinning. For Kolmi Op-Air face masks, Layer 1, 3 and 4 are consistent with PP, and Layer 2 and 5 are mostly consistent with PP by different manufacturing process. However the small peak at 9.3 and 34.7 degrees cannot be explained and may are caused by additional substances that give the mask its color, polarization properties or are needed in the manufacturing process. The measurements show that both face masks are made of the same material whether they come from a different company and have different colors. When comparing the layers of each graph in Figure 4 of supplemental file 6, it is obvious that the fourth peak is missing in 2 of the layers of each mask. This can be caused by differences in the manufacture process.

The three tests indicate that all 10 layers of each mask are made from the base material PP. Differences in melting temperature can be explained by the presence of a coating as indicated by the unexpected peaks as indicated in supplemental file 6, Figure 2a (Layer 4 and 5) and supplemental file 6, Figure 2b in the first DSC and in the XRD measurement. Also the differences in structure, shape, length and how fibers are interconnected, influence how fast the overheated steam penetrates the fibers and molecule structures [3]. Moreover, the glass transition temperature of fibrous PP is somewhere between 0 and -20 ^0^C depending on the ratio between atactic, syndiotactic and isotactic components [4,5]. Therefore, during the transitional state of 121 ^o^C, the level of crystallinity of the PP fiber as determined during fabrication in combination with internal material tension will result in differences in deformation resistance during sterilization.

Regarding H_2_O_2_ plasma sterilization it is likely that the low temperatures do not influence the properties of the PP filter material. However, the oxidation process most likely effect any surface ionization coatings on the fibers and therefore reducing the attraction of small particles during use.

**References**

[1]	Immirzi A, & Iannelli P. Whole-pattern approach to structure refinement problems of fibrous materials: application to isotactic polypropylene. Macromolecules 1988; 21(3), 768-773.

[2]	Cho D, Zhou H, Cho Y, Audus, D, & Joo Y. L. Structural properties and superhydrophobicity of electrospun polypropylene fibers from solution and melt. Polymer 2010; 51(25), 6005-6012.).

[3]	Mark H F. Encyclopedia of polymer science and technology, concise. John Wiley & Sons 2013.

[4]	Rodriguez-Arnold J, Zhang A, Cheng S.Z., Lovinger A. J., Hsieh E. T., Chu P., & Hawley, G.R. Crystallization, melting and morphology of syndiotactic polypropylene fractions: 1. Thermodynamic properties, overall crystallization and melting. Polymer 1994; 35(9), 1884-1895.

[5]	Janevski A, Bogoeva‐Gaceva G, & Mäder E. DSC analysis of crystallization and melting behavior of polypropylene in model composites with glass and poly (ethylene terephthalate) fibers. Journal of applied polymer science 1999; 74(2), 239-246.
