## Supplemental file 1 for "Sterilization of disposable face masks with respect to COVID-19 shortages; a nationwide field study including 19 sterilisation departments"

Supplemental file 1, DSC, XRD and FTIR tests on face mask materials

Differences in mask material was analysed by chemically and thermally comparing the fabric of the two most common types of face masks, showing different PFE after being sterilised once or twice with 121 ⁰C or H_2_O_2_ plasma sterilisation methods. A Differential Scanning Calorimetry (DSC), X-Ray Diffraction (XRD) and Transform InfraRed spectroscopy (FTIR) were conducted.

The chemical characterization test was conducted with a Fourier Transform InfraRed spectroscopy (FTIR) (Nicolet 6700 from ThermoFisher Inc., USA) to narrow down the potential polymer types that were used. Spectra was collected from 650 to 4000 cm^-1^ at a resolution of 2 cm^-1^ and averaged over 128 scans.

Thermal analysis of the sample material was conducted using Differential Scanning Calorimetry (DSC) calorimeter (PerkinElmer Diamond, <https://www.perkinelmer.com>), Figure 1, to investigate the thermal properties of the polymer layers. The method is based on the characteristic melting and crystallization transitions T_m_, which reflect the physical and chemical changes, endothermic and exothermic processes, or changes in heat capacity [20]. Samples of approximately 5 mg (figure 2) were cut and heated with a heating rate of 10 ⁰C/min. The temperature scan was set to -25 to 180 and 250 ⁰C and the cooling was conducted by nitrogen gas purge.

Additionally, the materials were tested with X-Ray Diffraction (XRD). XRD is a non-destructive test method used to analyse the structure of crystalline materials by identifying the crystalline phases present in a material and therefore reveals the chemical composition information. The experiments were conducted with a Bruker D8 Advance diffractometer (2theta-theta scan, often called Bragg-Brentano or focusing geometry) with Co Kα source (λ = 1.7889 Å, 35 kV and 40 mA) with Lynxeye position sensitive detector. The measurement range on a motorised varied divergent slit was set from 5 to 50 degrees with step size of 0.02 mm. A measuring time of 0.1 second per step was employed. Bruker software (DiffracSuite.EVA version 5.1, Bruker, USA) was used to interpret the data.

Furthermore an Infrared spectroscopy (FTIR) was conducted.

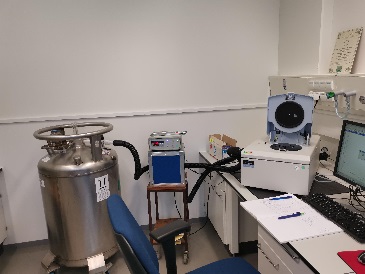

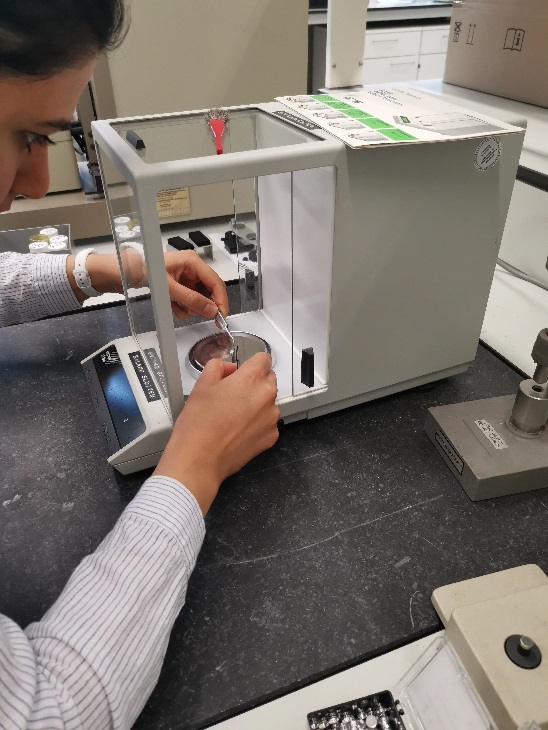

***Figure 1. Measurement setups. Left, Differential Scanning Calorimetry (DSC). Right,samples of 5 mg are accurately prepared for XRD measurements.***
