## Supplemental file 6 for "Sterilization of disposable face masks with respect to COVID-19 shortages; a nationwide field study including 19 sterilisation departments"

Supplemental file 6, Material analysis.

Differential Scanning Calorimetry (DSC), X-Ray Diffraction (XRD) and Transform InfraRed spectroscopy (FTIR) were conducted to determine the material properties in both the 3M Aura and Kolmi-Op-Air M52010. Figure 1 shows that 5 different polymeric layers of each mask consisting of different texture, fibre size and orientation as macroscopically observable.
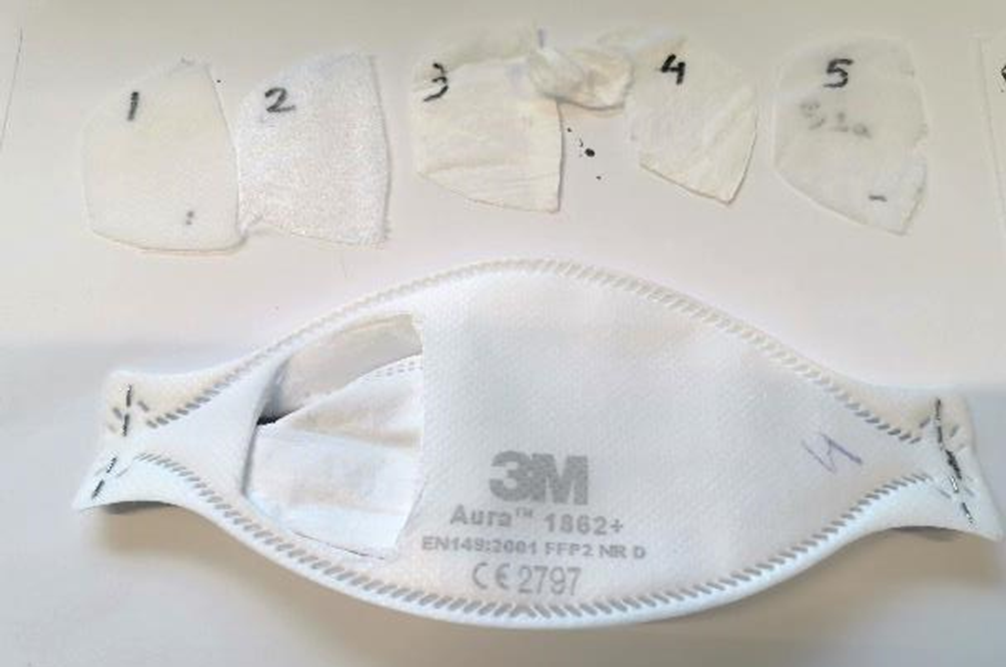

***Figure 1. Dissection of 5 constituent layers of 3M Aura 1862+ mask filter.***

They have individually undergone controlled thermal ramp for Differential Scanning Calorimetry. The first heating traces of each constituent layers are shown in Figure 2.

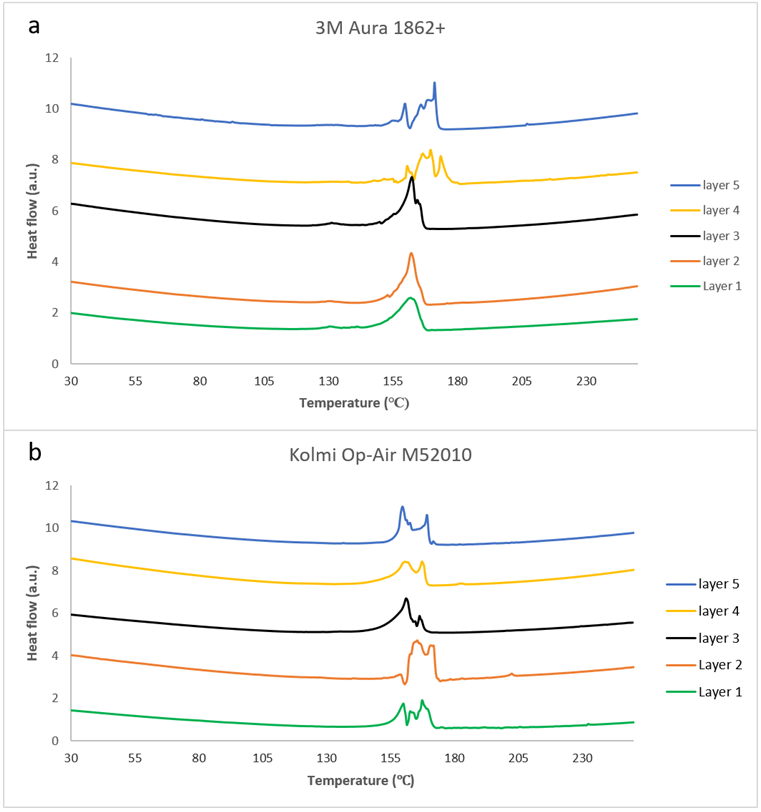
***Figure 2.* *DSC plots showing melting transitions of layers upon heating 10 degrees/min (endo up), a: 3M mask, b: Kolmi mask.***

Upon heating of each layer, broad endothermic melting transition occurred over a larger temperature range. In the 3M Aura 1862+ mask filter (Figure 2a), the melting transition of the three first layers occurred over 130-168 ℃ while the endpoints of layer 4 and 5 shift to 174℃ and 180℃ respectively. The melting transition peaks of the layers of Kolmi Op-Air M52010 masks varied in shape, and the melting occurred over 161-167 ℃ (Figure 2b). The enthalpy varied between 69 J/g for Layer 1 and 100 J/g for Layer 2 which indicate that these layers hold the same polymer composition or have a different fibre morphology, as the level of crystallinity is dependent on the polymer processing.

In order to eliminate the effect of fibre morphology and history of the samples, layer 1 has undergone a second heating following a cooling ramp after first heating. As shown in Figure 3 the serrated shape of the melting transition has switched to a smooth melting peak which occurs at exactly the same temperature.

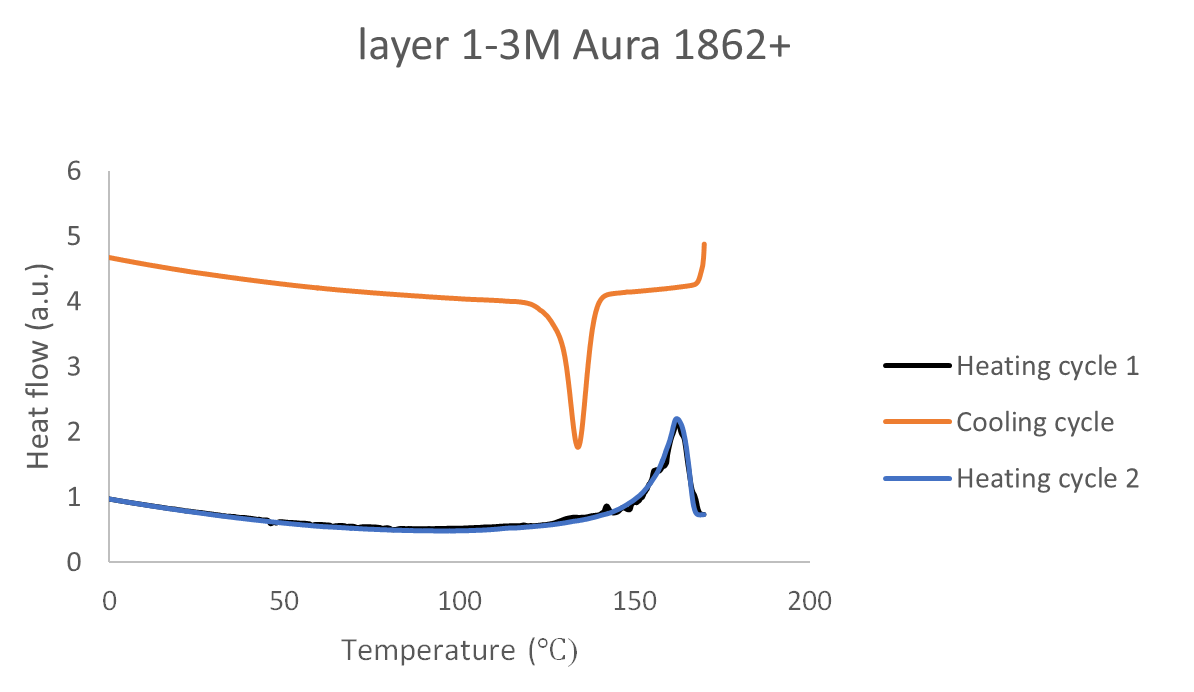

*Figure 3. DSC traces (heat-cool- heat cycles under 10℃/min) on layer 1 of 3M Aura 1862+ mask filter.*

This suggests that some of the DSC features in the first heating scan are due to (history dependent) artefacts, such as surface moisture, absorption of volatile substances. It is worth noting that normally DSC results on organic substances and polymers rely on using the second heating curve, as the first heating curve very frequently shows these artefacts. In the case of facemasks, and the actual use of the surface structure on the fibres in the mask material, we considered it useful to also show the first DSC heating curves.

*XRD measurements*

The Bruker D8 X-Ray Diffraction test results (Figure 4) for the 3M Aura 1862+ mask reveal 4 peaks for the 3M mask layers 1,2,3 and Kolmi 1,3,4 peaks at 16, 20, 22 and 25 degrees and 16, 20, 22 and 25 degrees.

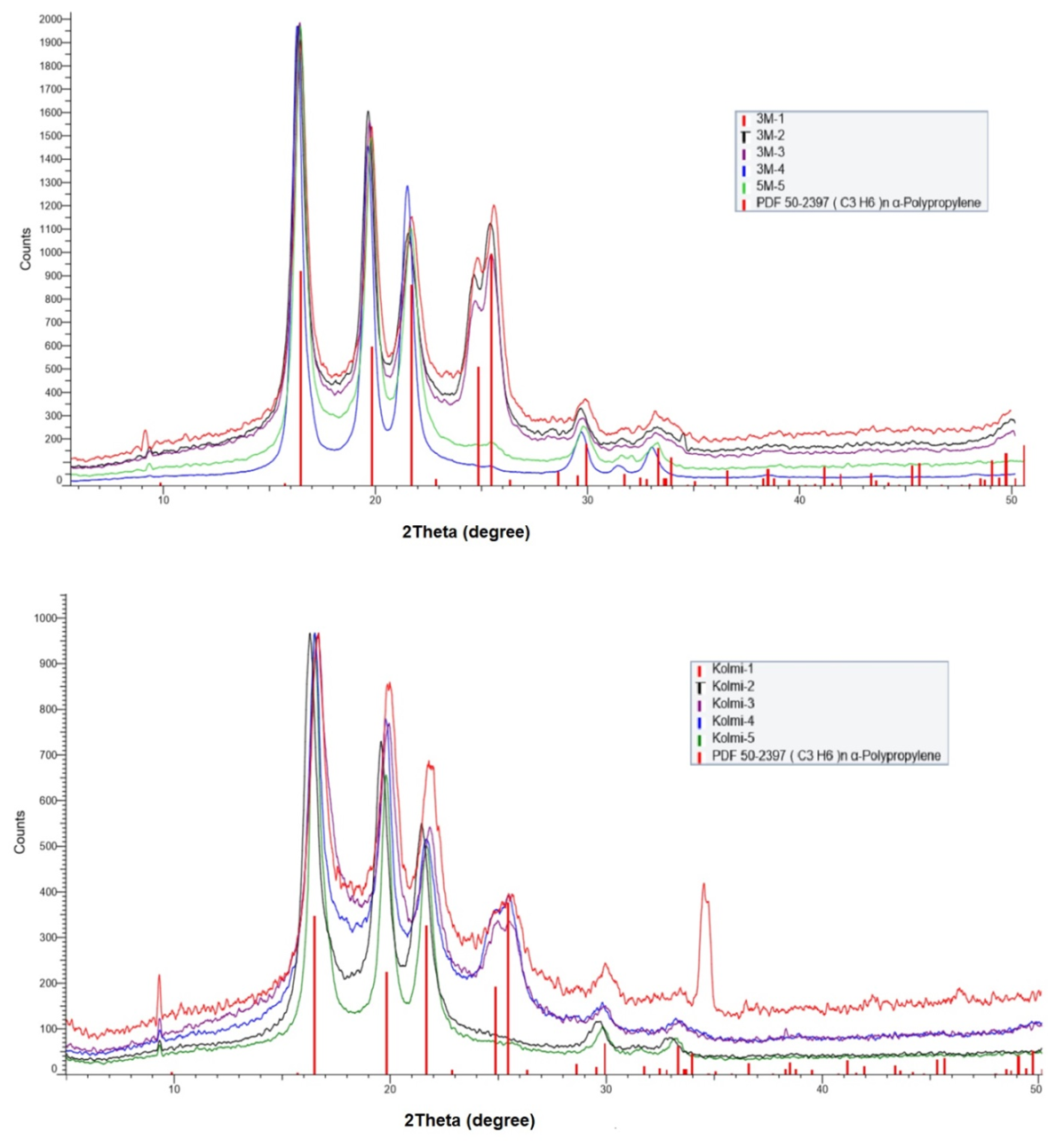

***Figure 4. The Bruker D8 X-Ray Diffraction test results with Polypropylene reference data for 3M (top) and Kolmi (bottom).***

For the layers 4 and 5 of the 3M mask, peaks were found at 16, 20 and 22 degree. For layers 2 and 5 of the Kolmi mask, peaks were also found at 16, 20 and 22 degrees. It was observed that the amplitude expressed in counts was higher for all layers of the 3M masks compared to the Kolmi.

*Infrared Spectroscopy (FTIR)*

Test results from the Infrared Spectroscopy confirmed a match of all 5 layers of both masks with the profile of the material PP. Figure 5 shows the comparison between layer 1 from 3M, layer 2 from Kolmi.

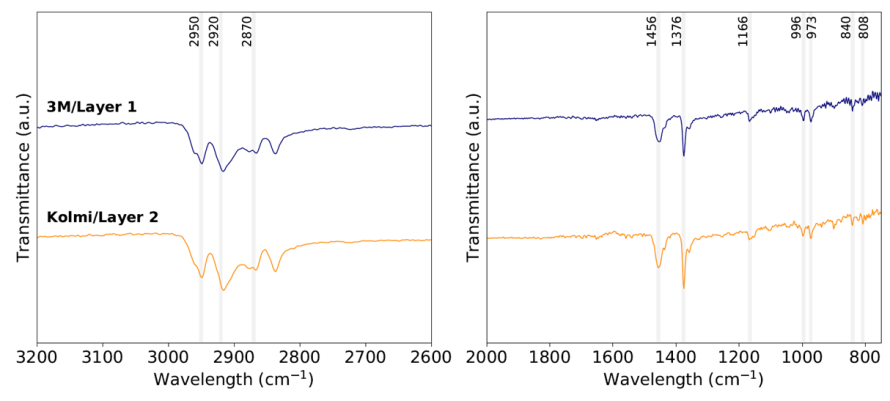

***Figure 5. The FTIR spectra of layer 1 from 3M mask (Top), and layer 2 from Kolmi mask (Bottom). Characteristics peaks of polypropylene are indicated by grey lines. The functional group assignment and vibration type of each PP peak can be found at Fang et al. (2012).***
